# Circulating insulin-like growth factor 1 (IGF-1) and short-term functional change in amyotrophic lateral sclerosis (ALS): exploratory evidence for systemic trophic-metabolic reserve

**DOI:** 10.64898/2026.11.05.26366903

**Authors:** Chai K Lim, Mariana Brocardo, Julie Hunter, Judith A. Heads, Dominic B Rowe, Sarah Furlong, Julie D Atkin

## Abstract

Amyotrophic lateral sclerosis (ALS) exhibit heterogeneous functional decline, potentially influenced by systemic metabolic reserve. We investigate whether circulating insulin-like growth factor 1 (IGF-1) is associated with subsequent functional change in ALS.

Serum IGF-1 was measured in 37 participants with ALS and 25 healthy controls. Baseline levels were compared between groups, and across disease-duration defined progression categories. Lagged regression assessed whether prior visit IGF-1 level predicted change in Revised ALS Functional Rating Scale (ALSFRS-R) on the next visit, adjusting for inter-visit interval, body mass index, age and sex. Mixed-effects and sensitivity analyses assessed robustness.

Baseline IGF-1 did not differ significantly between controls and ALS (mean ± SD: 107.3 ± 54.3 vs 132.2 ± 68.3 ng/mL, *p* = 0.12), or across progression categories (F(2, 34) = 0.13, p = 0.88). Across 48 intervals from 23 ALS participants, each 1-SD higher IGF-1 from prior visit was associated with 1.40-point less subsequent ΔALSFRS-R decline (95% CI: 0.03 to 2.77, p = 0.045). Generalised estimating equations yielded a similar association (β = 1.50, p = 0.030), whereas small-cluster robust inference showed wider uncertainty (p = 0.084). Repeated IGF-1 showed an intraclass correlation coefficient of 0.69, and the lagged association remained positive in all 23 leave-one-participant-out analyses. The IGF-1-by-time interaction was not significant in a random-slope mixed model (p =0.254).

Higher circulating IGF-1 may reflect systemic trophic-metabolic reserve associated with short-term functional preservation in ALS, but limited precision precludes definitive prognostic or causal conclusions.

## Introduction

Amyotrophic lateral sclerosis (ALS) is a rapidly progressing neurodegenerative disorder affecting motor neurons. Whilst its etiology is not fully understood, defects in energy metabolism are increasingly recognized as an important contributor to ALS pathophysiology, with a large body of evidence showing that motor neurons, skeletal muscle, and systemic metabolic networks all exhibit bioenergetic defects [1]. Identifying circulating biomarkers that reflect an individual’s capacity to withstand these metabolic and neurodegenerative stresses may therefore improve understanding of disease heterogeneity. Because ALS progression varies markedly between individuals, identifying what drives this variation is a critical step toward stratifying patients for prognosis and for precision-medicine trial design.

Insulin–like growth factor–1 (IGF-1) is a pleiotropic hormone involved in growth, energy homeostasis, skeletal muscle maintenance, mitochondrial function and cellular survival [2]. Lack of neurotrophic support is implicated in ALS and increasing evidence indicates that dysregulation of IGF-1 signaling contributes to pathogenesis [3, 4]. IGF-1 is protective in motor neurons [5] involving upregulation of mitophagy and protection of mitochondria from apoptosis [6]. Furthermore, IGF-1 is highly expressed in the relatively resistant oculomotor neurons in ALS [7]. These protective activities have led to therapeutic studies where beneficial effects of IGF-1 in both mouse models [5, 8–10] and clinical trials [11] have been observed. However, findings have not been consistent across experimental studies and clinical trials of recombinant IGF-1 have not demonstrated clear or reproducible therapeutic benefit in ALS [12–14]. For example, in a clinical trial for another neurodegenerative condition affecting motor neurons, spinobulbar muscular atrophy, no improvement in muscle strength or function was detected [15].

These conflicting findings suggest that the relationship between IGF-1 and neurodegeneration in ALS may be more complex than a simple linear association with the assumption that higher concentrations are uniformly beneficial. Heterogeneous factors such as age, nutritional status, endocrine regulation, receptor sensitivity and IGF-binding protein can affect IGF-1 signaling. Moreover, its effect can also differ between acute injury, chronic neurodegeneration and physiological aging. Consequently, endogenous circulating IGF-1 may represent an integrated indicator of systemic trophic and metabolic capacity rather than a simply direct measure of CNS IGF-1 activity or therapeutic target. Insufficient IGF-1 may indicate reduced trophic or metabolic reserve, whereas progressively higher concentration may provide insight into a plateau in therapeutic benefit once adequate signaling capacity is reached.

Consistent with this reserve-based framework, alterations in circulating IGF-1 have already been reported in ALS. IGF-1 is present in serum [16], and in ALS patients, lower serum IGF-1 concentrations have been detected compared to healthy controls [17]. In patients with long disease duration, circulating IGF-1 concentrations were also 36% higher than in controls [18]. These findings suggest that circulating IGF-1 may be associated with disease characteristics in ALS, although its clinical significance remains unclear. Moreover, most previous studies have relied on cross-sectional measurements, retrospectively estimated progression rates, or survival outcomes. It remains unclear whether IGF-1 measured at baseline is prospectively associated with functional change occurring before a subsequent visit.

In the present study, we examined circulating IGF-1 concentration in serum of individuals with ALS and healthy controls and evaluated whether IGF-1 was prospectively associated with subsequent functional change, measured using the revised ALS Functional Rating Scale (ALSFRS-R). The primary hypothesis was that higher IGF-1 at the preceding visit would be associated with less subsequent functional decline, after adjustment for age, sex, BMI, and the time interval between assessments. In addition, we explored whether the association between serum IGF-1 and ALS progression was linear, whether IGF-1 differed across clinical progression categories, and whether IGF-1 provided additional explanatory or classification information beyond clinical and trajectory-related measures. Because disease heterogeneity remains a key hurdle to biomarker development and precision medicine trial design in ALS, we tested the overarching hypothesis that serum IGF-1 captures systemic trophic-metabolic reserve contributing to functional resilience, rather than acting as a standalone biomarker of ALS progression.

## Methods

### **(A)** Study design and cohort

This study used a prospective longitudinal cohort design to examine disease heterogeneity. Participants were selected from the Macquarie University Motor Neuron Disease (MND) Research Centre’s Neurodegenerative Disease Biobank. All participants with ALS received a probable or definite diagnosis according to the revised El Escorial criteria [19], as determined by a consultant neurologist; eligible participants were aged 18 years or older and provided written informed consent or where the participant lacked capacity, consent via a legally authorised representative. Unaffected controls (hereafter referred to as healthy controls) without a diagnosis of ALS or another neurodegenerative disease were recruited in parallel through the same biobank; body mass index was not routinely collected for controls. Three highly heterogenous patient cohorts were carefully designed to stratify ALS patients into the desired subgroups while ensuring age and sex matching, regardless of stratification by subtype, disease progression or site of onset. Disease progression was based on the disease duration. Slow progression was defined as five years and over (≥ 5 years), normal disease progression was between two to five years (≥ 2 and < 5 years) and fast progression was defined as two years or less (< 2 years).

Analyses used different subsets of this cohort depending on data completeness: cross-sectional baseline comparisons required a single IGF-1 measurement per participant, the primary lagged analysis required paired consecutive visits with complete ALSFRS-R and BMI, and the mixed-effects trajectory analyses required complete ALSFRS-R and BMI at each individual visit. The derivation of each analytic sample is reported in the Results and summarised in the STROBE flow diagram (**Figure 1**).

**Figure 1.**
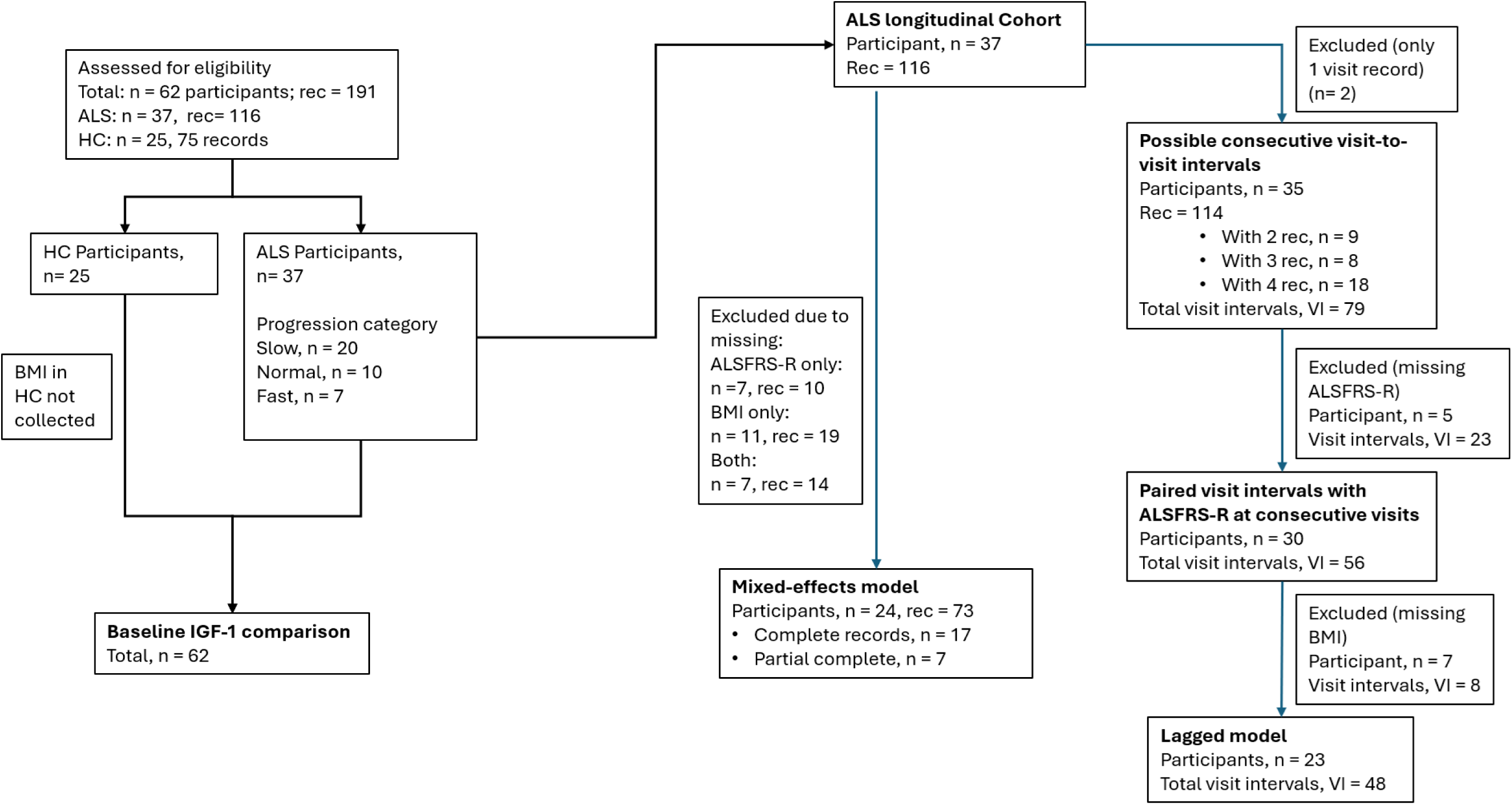
STROBE flow diagram of participant and visit-interval selection. All screened participants contributed to the cross-sectional baseline IGF-1 comparison; longitudinal analyses were progressively restricted to ALS participants with sufficient repeated visits and complete data, yielding separate analytic samples for the lagged and mixed-effects models. HC, healthy control; ALSFRS-R, Revised ALS Functional Rating Scale; BMI, body mass index; VI, visit-to-visit interval; rec, clinic visit record.

### **(B)** Laboratory analysis

#### Blood collection

Samples were collected and processed under standardised biobank procedures. Venous blood was collected by venipuncture (21G or 23G butterfly needle) into a 3.5 mL serum-separating tube (BD Vacutainer SST, cat. no. 367956; Becton Dickinson) drawn first in the order of draw and gently inverted per the manufacturer instructions. Samples were allowed to clot at room temperature for ≥30 min and centrifuged within 2 h of collection at 1,200 x *g* for 10 min at 20 °C with maximum brake. Serum was aliquoted into 500 µL volumes in barcoded 0.9 mL FluidX tubes and stored at −80 °C. Samples underwent no freeze-thaw cycles before analysis.

#### ELISA analysis of human serum

IGF-1 was quantified in serum using a sandwich enzyme-linked immunosorbent assay (ELISA, Cloud-Clone Corp ELISA Kit for IGF1, Catalogue CLC-SEA050Hu-5×96T). According to the manufacturer, the lowest limit of detection for IGF1 assay is around 58pg/ml and the inter-assay coefficient of variation, CV<12%

Briefly, a four-point standard curve was prepared in duplicate using components supplied by the kit, using a volume of 100μl and working concentrations ranging from 0.19ng/ml to 10ng/ml. The standards were then added to a 96 well plate, precoated with an antibody specific to IGF-1. 100μl of 1/50 diluted serum was also added to the plate in duplicate and incubated at 37°C for an hour. The serum was then removed, replaced with 100μl Detection Reagent A (biotin-conjugated antibody to IGF-1) and incubated for another hour at 37°C. After three washes with wash solution, 100µl of Detection Reagent B (Avidin conjugated to Horseradish Peroxidase (HRP) was added and incubated at 37°C for 30 mins.

The plate was then washed five times, before incubation with 90µl of TMB substrate for 10 – 20 minutes at 37°C in the dark. 50μl of stop solution was added. The plate was read at a wavelength of 450nm. All analyses were performed blinded.

### **(C)** Statistical Analysis

#### Study design and data structure

Analyses were conducted using a longitudinal dataset containing repeated Revised ALS Functional Rating Scale (ALSFRS-R) measurements and circulating serum IGF-1 concentrations. To avoid inflation from repeated measures in cross-sectional comparisons, baseline analyses were performed using the first available observation per participant. Longitudinal analyses were restricted to ALS participants and were conducted using consecutive visit-to-visit intervals.

Visit timing was recorded in days and converted to years for longitudinal modelling. For each participant (denoted i), consecutive visit-to-visit intervals were constructed. For each interval, the prior visit is denoted by (t-1) and the subsequent visit is denoted by (t). Lagged IGF-1 and lagged ALSFRS-R were defined as the values measured at visit (t-1) and the inter-visit interval was calculated as the time between visits (t-1) and (t), expressed in years. Short-term functional change (ΔALSFRS-R) was defined as:

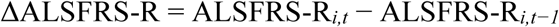

Negative values therefore indicate functional decline, whereas higher or less negative values (closer to 0) indicate less decline. Clinically meaningful decline was defined a priori as a decrease of at least 5 ALSFRS-R points between consecutive visits. Body mass index (BMI) was calculated from height and weight as kg/m² and then transformed using z-score normalization for multivariable models.

Progression categories were based on recorded disease duration and classified as slow, normal, or fast progression as described earlier. Because disease duration represented a minimum observed duration for participants who were alive or lost to follow-up, analyses involving progression categories were considered exploratory and descriptive.

#### Baseline group comparisons

Baseline IGF-1 concentrations were compared between ALS participants and healthy controls using Welch’s t-test to account for unequal variances. Effect size was quantified using Cohen’s d. Baseline IGF-1 concentrations were also compared across observed ALS progression categories using one-way analysis of variance (ANOVA).

#### Lagged regression analyses of visit-to-visit functional change with IGF-1

Lagged linear regression models were used to evaluate whether serum IGF-1 at prior clinic visit was associated with subsequent visit-to-visit change in ALSFRS-R. The primary outcome was ΔALSFRS-R, and the main predictor was lagged IGF-1 (standardized per SD). Models were adjusted for inter-visit interval, standardized baseline BMI, age, and sex using the following specification:

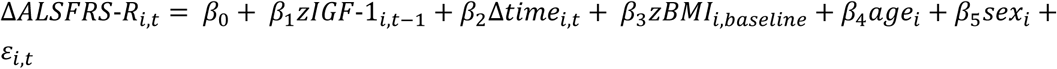

where zIGF-1_i,t-1_ denotes IGF-1 at the prior visit (standardized per SD); Δtime_i,t_ denotes the inter-visit interval in years; zBMI_i,baseline_ denotes baseline BMI (standardized per SD); age_i_ denotes age in years; sex_i_ denotes sex, with female as the reference group; and Ꜫ_i,t_ denotes the residual error.

Participant-level cluster-robust standard errors with normal-approximation inference were used for the primary lagged analysis to account for repeated visit-to-visit intervals within participants. To test robustness of the primary model, additional adjustment or sensitivity analyses were performed including (i) generalized estimating equations (GEE); (ii) conditional change models adjusted for ALSFRS-R at the prior clinic visit, denoted as ALSFRS-R*_i,t-1_*, to determine whether IGF-1 was associated with subsequent functional change independent of prior functional status; (iii) CR2 bias-reduced cluster-robust covariance estimation with Satterthwaite degrees of freedom as a conservative small-cluster estimate due to the modest sample size (n < 30).

#### Serum IGF-1 measurement variability and robustness analyses

A null random-intercept model for repeated IGF-1 measurements was used to estimate the intraclass correlation coefficient (ICC), grouping the observed variance into between-participant and within-participant plus residual components. To examine variation of IGF-1 levels coming from between-participant differences and visit-specific fluctuations, prior-visit IGF-1 measurement was separated into each participant’s mean concentration and the deviation of each measurement from that mean; both components were entered simultaneously in an adjusted GEE model. Robustness to individual participants was assessed by repeating the primary lagged analysis after excluding each participant in turn and summarizing the resulting IGF-1 coefficients.

#### Mixed-effects trajectory analyses

Mixed-effects models were used to examine whether baseline IGF-1 was associated with longitudinal ALSFRS-R trajectories. Fixed effects included time, baseline IGF-1 (standardized per SD), the IGF-1 × time interaction, baseline BMI, BMI x time, age and sex. Four models were fitted to the same complete-case sample: random-intercept models with and without IGF-1 terms, and random-intercept plus participant-specific random-slope models with and without IGF-1 terms. The random-intercept specification allowed participants to differ in baseline ALSFRS-R, whereas the random-slope specification additionally allowed the rate of ALSFRS-R change over time to vary between participants. Models used maximum-likelihood estimation for nested comparisons of fixed effects and random-effects structures; final coefficient estimates were obtained using restricted maximum likelihood.

Model fit was compared using Akaike information criterion (AIC), Bayesian information criterion (BIC), and likelihood-ratio tests. Nakagawa marginal and conditional R² were reported descriptively. Because participant-specific random slopes substantially improved fit, the random-slope model was given greater interpretive weight when assessing the baseline IGF-1 × time association.

#### Directional specificity

Reverse-direction models were fitted to examine whether prior functional status was associated with subsequent IGF-1 concentrations. This analysis was used to assess temporal (a)symmetry between IGF-1 and functional change as a follow-up to the lagged model. In this model, subsequent IGF-1 at visit *t*, standardized per SD, was modeled as the outcome and ALSFRS-R at prior visit (*t*−*1*) was the predictor of interest. The model was adjusted for standardized IGF-1 at prior visit (*t−1*) and relevant covariates with the following specification:

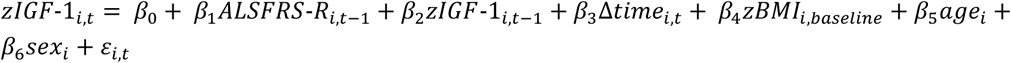

#### Exploratory analyses

Exploratory analyses of nonlinearity and candidate cutpoints, effect modification by observed progression category, incremental explanatory value, and binary decline classification were conducted as described in the Supplementary Methods. These analyses were considered exploratory for hypothesis-generating and were not considered for the current primary longitudinal conclusion.

#### Statistical Software

All analyses were conducted in R version 4.4.2 (R Foundation for Statistical Computing, Vienna, Austria). Data management was performed using dplyr, tidyr, and readr; mixed-effects models were fitted using lme4; conventional participant-clustered covariance estimates were obtained using sandwich; CR2 small-cluster corrections with Satterthwaite degrees of freedom were calculated using clubSandwich; generalized estimating equations were fitted using geepack; ridge-penalised logistic regression used glmnet; AUC analyses used pROC; and figures were generated using ggplot2. All statistical tests were two-tailed with statistical significance defined nominally as p < 0.05 for the prespecified primary normal approximation analysis. Results near this threshold were interpreted cautiously and further supported with sensitivity analyses given the modest number of participant clusters and the exploratory nature of secondary analyses.

## Results

### Study Cohort Demographics

The cohort comprised 37 individuals with ALS (25 males and 12 females) and 25 healthy controls (**Figure 1**). Baseline demographics and clinical characteristics are summarized in **Table 1**. Age and sex distribution did not differ significantly between ALS and control groups.

**Table 1.** Baseline & Longitudinal Characteristics of ALS and Control Participants.

|  | ALS | Controls | p-value |
|---|---|---|---|
| ***Baseline characteristics*** |  |  |  |
| Participants, n | 37 | 25 | - |
| Age, years, mean (SD) | 66.7 (8.2) | 65.3 (8.8) | 0.513<sup>1</sup> |
| Male/female, n; (male %) | 25/12 (67.6) | 18/7 (72) | 0.928<sup>2</sup> |
| BMI, kg/m<sup>2</sup>, mean (SD) | 26.3 (4.4) | - | - |
| ALSFRS-R, mean (SD) | 34.9 (8.0) | - | - |
| Time since diagnosis at baseline, years, median (IQR) | 0.5 (0.1 – 1.1) | - | - |
| Progression Categories |  |  |  |
| Disease Duration, years, median (IQR) | 5.91 (2.69 – 7.19) |  |  |
| Slow progressor, n (%) | 20 (54.1) | - | - |
| Normal progressor, n (%) | 10 (27.0) | - | - |
| Fast progressor, n (%) | 7 (18.9) | - | - |
| ***Longitudinal characteristics*** |  |  |  |
| Total visits, n | 116 | - | - |
| Total visit-to-visit intervals, n | 56 | - | - |
| Visits per participant, median (IQR) | 3 (2 – 4) | - | - |
| Follow-up duration (years), median (IQR) | 1.48 (0.69 – 2.28) | - | - |
| Inter-visit interval (years), median (IQR) | 0.50 (0.36 – 0.95) | - | - |
| ΔALSFRS-R per interval, median (IQR) | -4.0 (-6.25 – -1.00) | - | - |
| Annualised ALSFRS-R slope by progression category (points/year) |  |  |  |
| Slow progressor, n=16, mean (SD) | -3.78 (4.91) | - | - |
| Normal progressor, n=8, mean (SD) | -8.50 (6.13) | - | - |
| Fast progressor, n=7, mean (SD) | -15.50 (3.22) | - | - |

*Notes: Disease duration calculated as: Age at baseline − Age at diagnosis. Body mass index (BMI) is not available for controls. Amyotrophic Lateral Sclerosis Functional Rating Scale (ALSFRS-R) and progression categories apply only to ALS. Progression categories were calculated from disease duration with the following cutoff: ≥ 5 years = slow progressor; ≥2 and < 5 years = normal progressor; <2 years = fast progressor.*
*Statistical test: <sup>1</sup> Welch's t-test (continuous variables), <sup>2</sup> Chi-square test (categorical variables)*

Functional impairment was monitored using the ALSFRS-R. Across the longitudinal ALS group, participants had a median of 3 visits (IQR: 2 to 4) over a median follow-up duration of 1.48 years (IQR: 0.69–2.28), with a median inter-visit interval of 0.5 years (IQR: 0.36– 0.95) (**Figure 1**). Across all available visit-to-visit intervals, the median change in ALSFRS-R was -4.00 points (IQR: -6.25 – -1.00), indicating measurable functional decline between clinic visits.

Of 37 ALS participants, 31 had at least two ALSFRS-R assessments, with a median annualised ALSFRS-R decline of 5.2 points per year (IQR: 2.7–13.5). Participant-level annualized slopes for ALSFRS-R differed markedly across three observed progression categories: (a) Slow: mean = -3.78 points/year, (b) normal: mean = -8.5 points/year, (c) Fast: mean = -15.5 points/year (**Table 1**). One-way ANOVA confirmed significant differences between these categories (F(2, 28) = 13.80, p < 0.001, η² = 0.50). Post-hoc comparisons demonstrated significant separation between slow and fast progressors (Bonferroni-adjusted p < 0.001), borderline separation between normal and fast progressors (p = 0.051), and no significant difference (p = 0.248) between slow and normal groups. These findings confirm that the observed progression categories captured substantial heterogeneity in longitudinal ALSFRS-R decline.

### Baseline IGF-1 Levels in ALS and Controls

Baseline IGF-1 concentrations were compared between ALS participants (n = 37) and healthy controls (n = 25) cross-sectionally. Mean serum IGF-1 was higher in ALS (132.2 ± 68.3) than in controls (107.3 ± 54.3), corresponding to a mean difference of 24.9 ng/ml and a moderate effect size (Cohen’s d = 0.40). However, this difference did not reach statistical significance (Welch t-test, p = 0.12; **Figure 2**).

**Figure 2.**
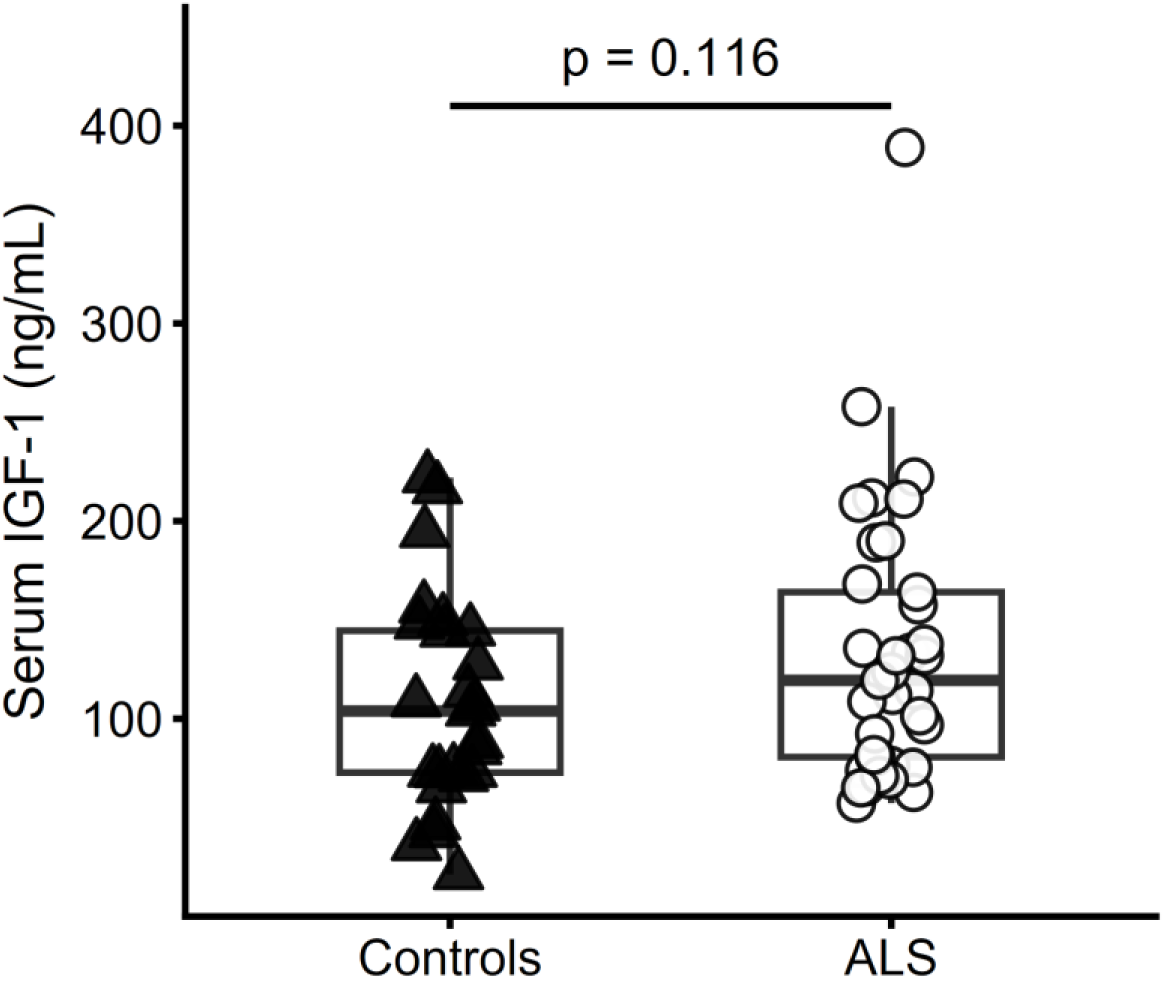
Baseline IGF-1 concentrations in ALS participants and healthy controls. Boxplots represent the median and interquartile range. Group differences were assessed using Welch’s t-test with p-value denoted by p presented.

### Baseline IGF-1 Levels across observed ALS Progression Categories

Baseline serum IGF-1 concentrations were cross-sectionally compared across observed ALS progression categories based on disease duration. Baseline serum IGF-1 did not differ between slow, normal and fast progressors (F(2, 34) = 0.13, p = 0.882, **Figure 3**), although fast progressors showed the lowest circulating IGF-1 levels.

**Figure 3.**
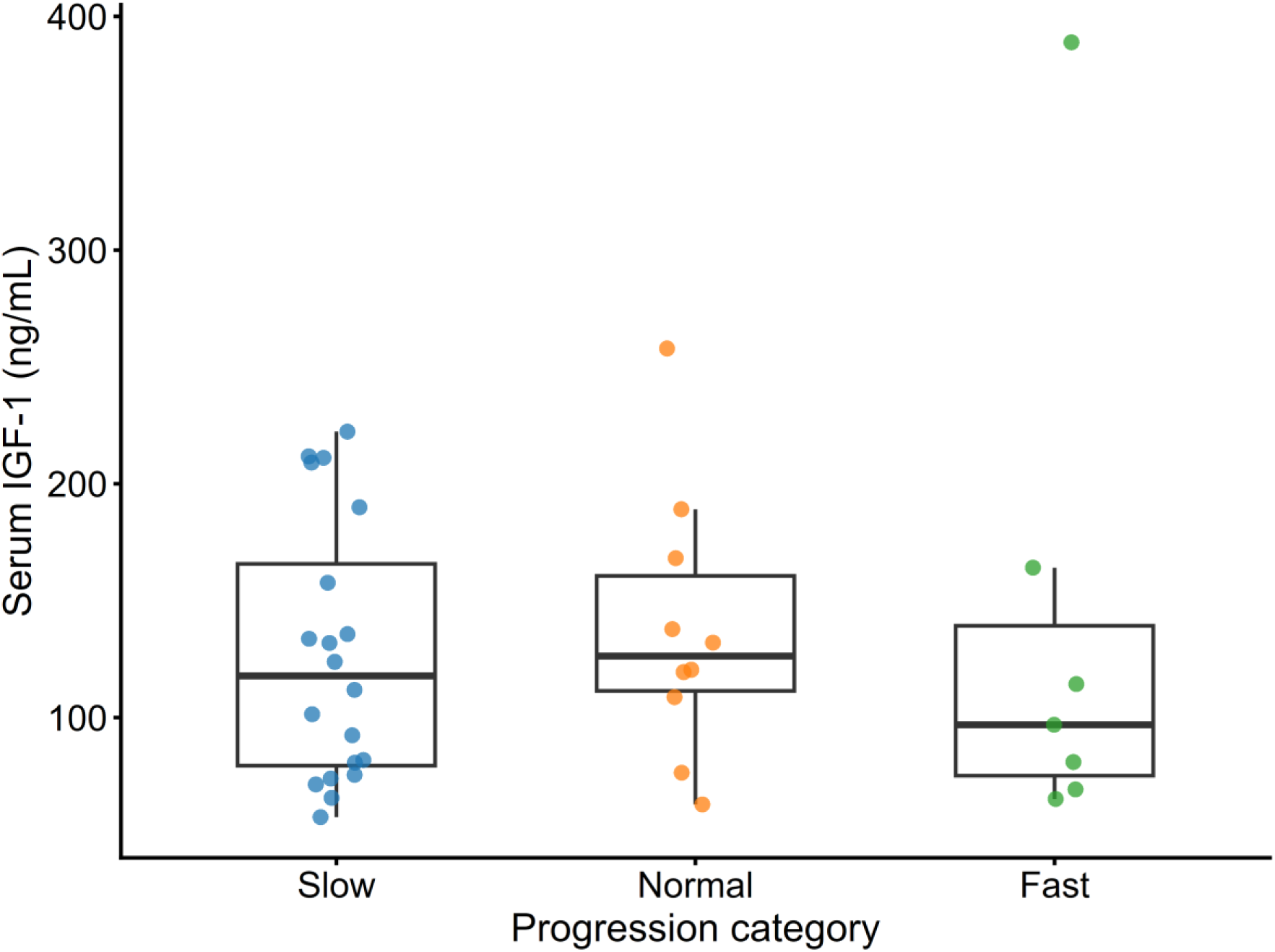
Baseline serum IGF-1 concentrations across observed ALS progression categories. Boxplot represent the median and interquartile range of IGF-1 concentrations in slow (blue dots), normal (orange dots) and fast (green dots) progression. Group differences were assessed using one-way ANOVA showing no differences across all groups.

### Prospective Association Between IGF-1 and Short-Term Functional Change

We next examined whether serum IGF-1 concentrations at prior clinic visit were associated with subsequent visit-to-visit change in ALSFRS-R (**Table 2**). The complete case analysis included 48 intervals from 23 ALS participants (**Figure 1**). In the primary model adjusted for inter-visit interval, baseline BMI, age and sex, every 1 SD higher IGF-1 concentration at the prior visit was associated with a 1.40-point higher subsequent ΔALSFRS-R (95% CI 0.03 to 2.77, p = 0.045), implying less functional decline at the next follow-up visit.

**Table 2.** Primary lagged models examining the association between prior-visit IGF-1 and subsequent ALSFRS-R change.

| **Model** | **n** | **β estimate (95% CI)** | **p-value** | **Covariates adjusted** |
|---|---|---|---|---|
| Lagged linear model, Cluster-robust SE | 48 intervals, n = 23 | 1.40 (0.03, 2.77) | 0.045 | Inter-visit interval, BMI, age, sex |
| GEE model as sensitivity analysis | 48 intervals, n = 23 | 1.50 (0.15, 2.86) | 0.030 | Inter-visit interval, BMI, age, sex |
| Conditional change model (adjust for prior ALSFRS-R) | 48 intervals, n = 23 | 1.45 (0.10, 2.80) | 0.036 | Inter-visit interval, prior ALSFRS-R, BMI, age, sex |
| Lagged linear model adjusted for small-cluster t-based inference | 48 intervals, n = 23 | 1.40 (-0.22, 3.02) | 0.084 | Inter-visit interval, BMI, age, sex |
| Conditional change model adjusted for Small-cluster t-based inference | 48 intervals, n = 23 | 1.45 (-0.15, 3.05) | 0.071 | Inter-visit interval, prior ALSFRS-R, BMI, age, sex |

*The outcome was short-term functional change, defined as ΔALSFRS-R = ALSFRS-R at the subsequent visit minus ALSFRS-R at the prior visit. Therefore, higher or less negative ΔALSFRS-R values indicate less functional decline. IGF-1 and baseline BMI were standardised per SD. β values represent the difference in subsequent ΔALSFRS-R associated with a 1-SD higher IGF-1 concentration at the prior visit. Age was entered in years, and female was the reference category for sex. For the lagged linear and conditional change models, participant-level cluster-robust SE with normal-approximation inference were used to account for repeated visit-to-visit transitions within participants. The GEE model was fitted as a sensitivity analysis. Small-cluster t-based inference using CR2 cluster-robust covariance estimation and Satterthwaite degrees of freedom was examined because the number of participant clusters was modest. ALSFRS-R = revised ALS Functional Rating Scale; BMI = body mass index; CI = confidence interval; CR2 = bias-reduced cluster-robust covariance estimator; GEE = generalised estimating equations; SE = standard error; SD = standard deviation.*

The estimate was similar after additional adjustment for prior ALSFRS-R (β = 1.45, 95% CI 0.10 to 2.80; p = 0.036) in the conditional change model and in the GEE sensitivity analysis (β = 1.50, 95% CI 0.15 to 2.86; p = 0.030). Under conservative CR2/Satterthwaite small-cluster inference, the corresponding effect estimates remained similar in magnitude and direction, but confidence intervals widened and included zero for both the lagged model (β = 1.40, 95% CI -0.22 to 3.02; p = 0.084) and the conditional model (β = 1.45, 95% CI -0.15 to 3.05; p = 0.071). Thus, the estimated association was consistent across specifications, although its precision was sensitive to inference with the modest sample size.

### Stability and source of variation in repeated IGF-1 measurement

Repeated IGF-1 measurements showed an ICC of 0.69, indicating that approximately 69% of observed variability occurred between participants, whereas the remaining 31% reflected within-participant variation over time, together with residual measurement variability. In the within-versus between-participant comparison, participants with a higher average IGF-1 concentration showed more preserved subsequent function (between-participant β = 1.67, 95% CI 0.14 to 3.21; p = 0.032). By contrast, visit-specific deviations from a participant’s usual IGF-1 level were not associated with subsequent functional change (within-participant β = -0.01, 95% CI -1.00 to 0.98; p = 0.989).

In leave-one-participant-out analyses, the lagged IGF-1 coefficient remained positive after each of the 23 participants was excluded in turn. Estimates ranged from 0.95 to 1.86, with a median of 1.44, close to the full-sample estimate of 1.40. The direction and approximate magnitude of the lagged association were therefore not driven by any single participant.

### Baseline IGF-1 and longitudinal ALSFRS-R trajectories

We next examined whether baseline IGF-1 was associated with longitudinal ALSFRS-R trajectories using mixed-effect models (**Figure 4**). In the random-intercept model, the IGF-1 × time interaction was positive and statistically significant (β = 2.54 ALSFRS-R points/year per SD; p = 0.0006). Adding baseline IGF-1 and the IGF-1 × time interaction improved fit relative to the corresponding model without IGF-1 (model 1 vs model 2: ΔAIC = -7.19; ΔBIC = -2.61; likelihood-ratio χ²(2) = 11.19, p = 0.0037) and increased marginal and conditional R² by 0.051 and 0.062, respectively (**Table 3**).

**Figure 4.**
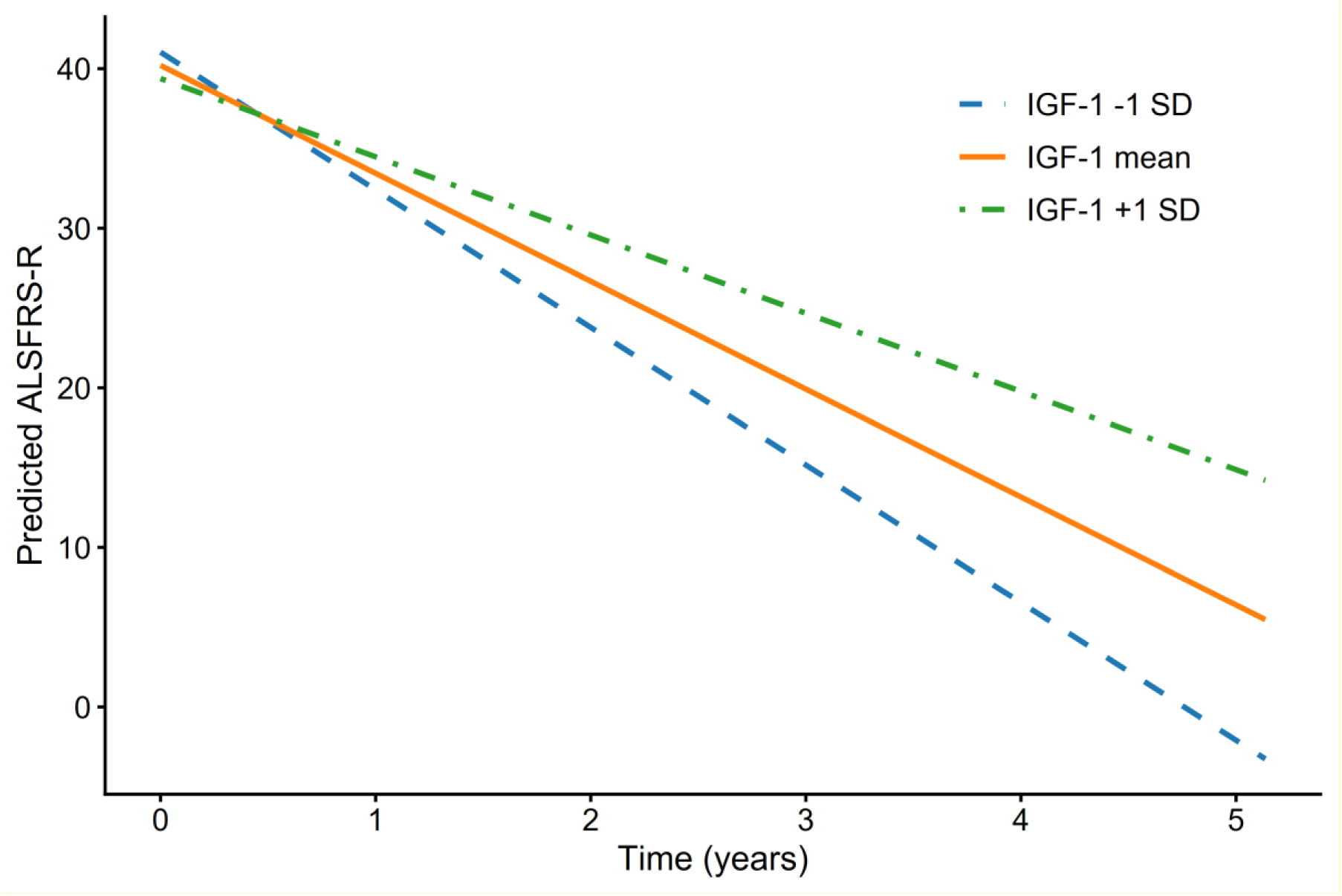
Predicted ALSFRS-R trajectories by baseline IGF-1. Population-level predictions from the random-slope mixed-effects model are shown for baseline IGF-1 concentrations at -1 SD, the mean, and +1 SD, with age, sex, and baseline BMI held constant. Higher baseline IGF-1 corresponded to numerically slower predicted decline, although the IGF-1 × time interaction was less precisely estimated in the better-fitting random-slope model compared to other models.

**Table 3.** Comparison of linear mixed-effects models of longitudinal ALSFRS-R trajectories.

| Predictor | Model 1:<br>Base<br>(RI) | Model 2:<br>Base + IGF-1<br>(RI) | Model 3:<br>Base<br>(RI + RS) | Model 4:<br>Base + IGF-1<br>(RI + RS) |
|---|---|---|---|---|
| **Fixed Effects** |  |  |  |  |
| (Intercept) | **45.92 (9.87)** | **47.52 (10.28)** | **46.30 (8.62)** | **45.73 (9.02)** |
| Time, years | **-4.28 (0.61)** | **-5.42 (0.63)** | **-7.04 (1.15)** | **-6.76 (1.15)** |
| Baseline IGF-1, per SD | — | -1.31 (2.01) | — | -0.83 (1.70) |
| Baseline BMI, per SD | 0.39 (1.38) | 0.03 (1.53) | 0.40 (1.13) | 0.18 (1.27) |
| Age, years | -0.14 (0.15) | -0.16 (0.15) | -0.13 (0.13) | -0.12 (0.14) |
| Male sex (ref =female) | 2.16 (2.64) | 2.33 (2.72) | 2.55 (2.34) | 2.53 (2.43) |
| Baseline IGF-1 x time | — | **2.54 (0.74)** | — | 1.86 (1.63) |
| Baseline BMI x time | -1.30 (0.80) | -0.69 (0.75) | -1.25 (1.14) | -0.79 (1.20) |
| **Random Effects** |  |  |  |  |
| Residual | 17.42 | 14.09 | 4.11 | 4.09 |
| Intercept: Participant | 31.02 | 34.54 | 24.87 | 26.99 |
| Slope: Time | — | — | 23.59 | 22.72 |
| Intercept-Slope Correlation | — | — | 0.107 | 0.094 |
| **Model Fit Statistics** |  |  |  |  |
| AIC | 469.32 | 462.14 | 439.63 | 441.97 |
| BIC | 487.65 | 485.04 | 462.54 | 469.46 |
| Marginal R<sup>2</sup> | 0.3420 | 0.3934 | 0.453 | 0.408 |
| Conditional R<sup>2</sup> | 0.7347 | 0.7964 | 0.963 | 0.957 |
| **Nested Model Comparisons** |  |  |  |  |
| Model 1 vs Model 2 | (Reference) | **ΔAIC = −7.19; p=0.004** | — | — |
| Model 3 vs Model 4 | — | — | (Reference) | ΔAIC = 2.34; p=0.435 |
| Model 2 vs Model 4 | — | (Reference) | — | **ΔAIC = −20.17** |

*Notes: All models used the same complete-case sample of 73 observations from 24 participants. Fixed-effect estimates, standard errors, p-values, and random-effect components were obtained from models fitted using restricted maximum likelihood. AIC, BIC, and likelihood-ratio tests were obtained from models fitted using maximum likelihood. The base model (model 1) covariate set comprised time in years, standardized baseline BMI, baseline BMI × time, age in years, and sex. Models containing IGF-1 additionally included standardized baseline IGF-1 and baseline IGF-1 × time. RI, random intercept; RS, participant-specific random slope for time. Marginal R² represents variance explained by fixed effects; conditional R² represents variance explained by fixed and random effects combined. Negative ΔAIC and ΔBIC favor the expanded model. The likelihood-ratio tests for adding IGF-1 jointly evaluate the baseline IGF-1 main effect and IGF-1 × time interaction. Sex is shown as male relative to the female reference category. Baseline IGF-1 and BMI are expressed per standard deviation (SD). Standard errors are presented in parentheses (SE).* ***Bold:*** *p < 0.001 unless otherwise specified.*

Since the rate of ALSFRS-R decline varied substantially between individuals, we also fitted a model allowing participant-specific random slopes for time (model 3). This random-slope model improved overall model fit compared with the random-intercept IGF-1 model (model 2 vs model 4: ΔAIC = −20.17; ΔBIC = -15.58; likelihood-ratio χ²(2) = 24.17, p = 5.66 x 10^-6^), demonstrating marked between-participant heterogeneity in rates of functional decline. Using this better-fitting random-slope structure, adding IGF-1 and the IGF-1 × time interaction did not improve model fit (model 3 vs model 4: ΔAIC = 2.34; ΔBIC = 6.92; likelihood-ratio χ²(2) = 1.66, p = 0.435). Nonetheless, the IGF-1 × time estimate remained positive but was estimated with considerable uncertainty (β = 1.86 ALSFRS-R points/year per SD, 95% CI -1.34 to 5.06; p = 0.254).

Taken together, the direction of the mixed-effects estimates was consistent with the lagged-model findings, with higher baseline IGF-1 corresponding to slower functional decline in the random-intercept model. When participant-specific random slopes were included, substantially improving model fit, the IGF-1 × time estimate remained positive but was attenuated, estimated less precisely, and was not statistically significant. These findings indicate substantial between-participant heterogeneity in progression rates and limited precision in the estimated association between baseline IGF-1 and long-term ALSFRS-R trajectories.

### Directional Specificity

To clarify the relationship between ALSFRS-R and IGF-1 concentration, we further asked whether prior ALSFRS-R predicted subsequent IGF-1 level, testing bidirectionality relative to the primary lagged model. To examine this reverse temporal direction, we modeled subsequent IGF-1 as the outcome and prior ALSFRS-R as the predictor, adjusting for prior IGF-1 and relevant covariates. Prior ALSFRS-R was not significantly associated with later IGF-1 concentrations (p = 0.096), whereas prior IGF-1 strongly predicted subsequent IGF-1 (p < 0.001). Because the reverse-direction estimate was non-significant rather than clearly null and the sample was modest, these findings are consistent with, but do not establish, temporal asymmetry.

### Exploratory analyses

Exploratory spline modeling suggested a possible nonlinear pattern, with lower prior-visit IGF-1 concentrations corresponding to greater subsequent functional decline and apparent flattening at higher concentrations (**Figure S1**). However, the overall spline terms were not statistically significant (p = 0.19). Other analyses, including modeling of clinically meaningful cutpoints, effect modification across observed progression categories, and participant-grouped classification of subsequent decline, were performed to explore the possible clinical utility of IGF-1, but none were conclusive given the modest sample size. These data may inform the design of future, adequately powered studies. Full methods and results are provided in the Supplementary Materials.

## Discussion

This longitudinal study found that higher serum IGF-1 concentrations at the prior visit were associated with more preserved short-term functional change in Australian ALS patients. In the lagged model, each 1 SD higher IGF-1 level at the prior visit corresponded to ∼1.4 fewer points of functional loss over the subsequent visit while accounting for BMI, age, sex, and inter-visit interval. The magnitude remained similar even after several sensitivity analyses. More conservative analysis to account for the modest sample size produced nearly identical point estimates but less precise confidence intervals, indicating our finding was consistent, but the precision remains uncertain and further replication in a larger cohort is needed. A previous cross-sectional study showed that IGF-1 concentrations correlated with disease severity assessed by ALSFRS-R [16]. However, our findings add new insight that IGF-1 is not merely a synchronous correlate of disease severity but is linked to subsequent trajectory that can only be addressed in a longitudinal study design. Because circulating IGF-1 levels vary between individuals, and analytical variability can further confound the findings, several post-data analyses were performed to clarify the stability and likely source of heterogeneity in IGF-1 levels. Repeated IGF-1 measurements were relatively stable within individuals (ICC = 0.69), and the between-participant component, rather than short-term deviations from an individual’s usual IGF-1 concentration, was associated with subsequent functional change. The lagged coefficient also remained positive in every leave-one-participant-out analysis and stayed close to the full-sample estimate. Together, these findings argue against the result being driven by a single influential participant and suggest that IGF-1 may operate primarily as a relatively stable participant-level marker. The reverse-direction analysis did not support evidence of prior ALSFRS-R in predicting subsequent IGF-1 concentrations, suggesting temporal asymmetry between IGF-1 and functional change in ALS progression, although it was not sufficiently precise to establish directionality or causation.

Mixed-effects modelling further clarified this relationship. In the random-intercept model, we examined whether participants with different starting IGF-1 levels also differed in their ALSFRS-R scores at the start of follow-up, whereas the IGF-1 × time interaction examined whether they subsequently declined at different rates. The IGF-1 × time interaction was statistically significant, indicating that participants with higher baseline IGF-1 showed slower longitudinal decline. However, allowing participants to have individual progression rates substantially improved model fit. In this more flexible and better-fitting model, the association between starting IGF-1 levels and subsequent rates of decline remained positive but was attenuated, estimated less precisely, and was not statistically significant (p = 0.254). The central findings therefore rest on the lagged visit-to-visit analyses rather than on the longitudinal trajectory models. Taken together, higher IGF-1 may be associated with slower average functional decline, but baseline IGF-1 did not clearly account for the substantial heterogeneity in longer-term ALS progression rates.

While previous studies have examined associations between circulating IGF-1 and ALS survival or overall prognosis, the present study extends the field by using repeated biomarker measurements and lagged longitudinal analyses to investigate temporal relationships between IGF-1 and functional decline. The higher IGF-1 levels predicted better preservation of ALSFRS-R scores during the subsequent assessment interval, whereas prior functional status did not predict later IGF-1 levels. This temporal asymmetry, together with the consistency of findings across sensitivity analyses, suggests that circulating IGF-1 may reflect a component of systemic trophic-metabolic reserve that precedes and potentially influences short-term functional trajectories in ALS, rather than simply mirroring disease severity.

### IGF-1 Signaling and Motor Neuron Survival

These statistical findings raise the question of which biological mechanism may plausibly link higher circulating IGF-1 to preserved short-term function in ALS. IGF-1 is a pleiotropic neurotrophic and metabolic factor that activates the PI3K–Akt and MAPK signaling pathways, promoting neuronal survival, axonal growth, synaptic maintenance and cellular stress responses [2]. In motor neurons, IGF-1 signaling has been linked to anti-apoptotic cascades, stabilizes mitochondrial function, and supported axonal transport [6]. IGF-1-related pathways may also contribute to non-cell autonomous mechanisms in ALS. For example, modulation of IGF1R-mTOR pathway in mutant SOD1 astrocytes has been reported to be protective against motor neuron toxicity [20]. Moreover, IGF-1 can delay motor neuron degeneration and prolong survival in some ALS mouse models [5], although findings have not been consistent across all experimental studies [13].

The prospective association observed here is biologically coherent with this framework. Higher circulating IGF-1 may reflect more preserved trophic signaling capacity, greater systemic anabolic reserve, or more intact downstream pathway responsiveness. Even modest preservation of these signaling mechanisms could translate into measurable short-term differences in functional decline, particularly in a disease characterized by cumulative motor neuron loss.

Exploratory supplementary analyses suggested a possible nonlinear pattern, with visually greater subsequent decline at lower IGF-1 concentrations and a possible flattening of the association at higher concentrations. However, the overall spline test was not statistically significant, and the tertile, threshold, and hinge analyses did not identify a robust cutpoint. These findings therefore do not establish either a specific functional form or a clinically meaningful IGF-1 threshold and should be regarded as hypothesis-generating. Larger studies with adequate power are needed to determine whether the association is linear, nonlinear, or characterized by a biologically relevant threshold.

### Compensatory Upregulation and Disease Biology

ALS participants in this Australian cohort exhibited numerically higher serum IGF-1 concentrations compared to healthy controls, although the difference was not statistically significant. This finding is consistent with previous studies reporting no clear difference in serum IGF-1 between ALS participants and controls [16, 18, 21]. One possible interpretation is that peripheral IGF-1 may increase as part of a compensatory response to neurodegeneration, muscle loss, or systemic metabolic stress. However, higher peripheral IGF-1 does not necessarily indicate more effective IGF-1 signaling within the CNS. Receptor-level resistance, altered post-receptor signaling, changes in IGF-binding proteins, or impaired transport across the blood-brain barrier may all limit the functional aspects of circulating IGF-1.

Within the ALS group, relatively higher serum IGF-1 may therefore reflect either more effective trophic compensation for neuronal damage, or a broader marker of preserved systemic metabolic and anabolic capacity. This prospective association between higher IGF-1 and attenuated short-term functional decline, even after adjusting for age, sex, BMI, and prior ALSFRS-R, is consistent with this interpretation. However, the present observational data cannot delineate whether IGF-1 is directly contributing to neuroprotection or instead acting as a marker of preserved systemic physiological resilience.

### IGF-1 and Metabolic Reserve

ALS is increasingly recognized as a multisystem disorder involving metabolic alterations, including hypermetabolism, altered glucose utilization, skeletal muscle dysfunction and weight loss [1]. As a regulator of anabolic signaling and skeletal muscle maintenance, IGF-1 may capture aspects of this systemic metabolic state that are not fully reflected by BMI alone. Although BMI was included as a covariate in the longitudinal models, BMI provides only a broad measure of an individual’s metabolic status and does not capture muscle mass, endocrine function, or anabolic capacity.

From a resilience perspective, serum IGF-1 may indicate one component of systemic trophic-metabolic reserve. Individuals with more preserved anabolic and metabolic signaling may be better able to withstand the energetic demands imposed by ongoing neurodegeneration and muscle wasting. This aligns with the prospective association between higher IGF-1 and more preserved short-term function, while also recognizing that IGF-1 provided only limited incremental explanatory information in exploratory linear models and did not improve either apparent or participant-grouped cross-validated classification of clinically meaningful decline. Exploratory supplementary analyses were compatible with a possible flattening of the association at higher concentrations but did not statistically establish a nonlinear or plateauing relationship.

### Biomarker Positioning

Taken together, these findings position serum IGF-1 as a biologically plausible and temporally coherent marker associated with short-term functional change in ALS, but not as a standalone prognostic biomarker. Higher IGF-1 was prospectively associated with more preserved short-term ALSFRS-R change. However, baseline IGF-1 concentrations did not differ across the observed slow, normal, and fast progression categories, and there was no evidence that the association between lagged IGF-1 and subsequent functional change varied across these groups. IGF-1 also did not improve internal classification of clinically meaningful decline. These findings suggest that circulating IGF-1 may capture a trophic-metabolic signal associated with short-term functional preservation rather than clearly distinguishing broader patterns of disease progression.

IGF-1 may therefore reflect one component of trophic-metabolic resilience rather than the full complexity of ALS progression. This distinction is important because ALS progression reflects multiple converging pathobiological processes, including excitotoxicity, mitochondrial dysfunction, RNA dysregulation, protein aggregation, neuroinflammation, skeletal muscle dysfunction and systemic metabolic stress [22]. A single circulating trophic marker is therefore unlikely to capture the overall disease trajectory.

### Implications for Therapeutic Targeting

The findings have implications for therapeutic strategies targeting the IGF-1 pathway. Previous trials of exogenous IGF-1 administration in ALS did not demonstrate clear or consistent clinical benefit, although these trials were considered to have a high risk of bias [14]. The present results do not contradict those trial findings. Rather, they suggest that endogenous circulating IGF-1 may be associated with short-term functional preservation, potentially as a marker of broader trophic-metabolic reserve.

This distinction is important because an observational association with endogenous IGF-1 does not necessarily imply that pharmacologically increasing IGF-1 will slow ALS progression. Circulating IGF-1 may act as a marker of preserved physiological reserve rather than a direct modifiable therapeutic target or outcome measure in ALS. Future studies should therefore examine not only circulating IGF-1 concentrations, but also downstream pathway activation such as Akt phosphorylation, IGF-1 binding protein dynamics, interaction with other metabolic markers, and longitudinal functional outcomes.

### Limitations

This study has several limitations. Although the parent study had a prespecified sample size for its original objectives, missing measurements and the requirement for complete data reduced the effective sample available for the present longitudinal analyses. The primary lagged and mixed-effects analyses were both based on modest complete case samples (**Figure 1**). The modest number of participant clusters limited statistical precision: although the primary lagged association was statistically significant using participant-clustered normal-approximation inference and remained directionally consistent in leave-one-participant-out analyses, its confidence interval widened under the more conservative CR2 small-cluster correction. The study also lacked external validation and did not include direct mechanistic measures such as muscle mass, nutritional biomarkers, IGF-binding proteins, or downstream pathway activation. Replication in larger longitudinal cohorts with more comprehensive metabolic and mechanistic profiling is therefore required.

## Conclusion

Higher circulating IGF-1 was prospectively associated with more preserved short-term functional change in ALS after adjustment for age, sex, standardized baseline BMI, and inter-visit interval, and the association remained present in a conditional model additionally accounting for prior functional status. However, uncertainty increased under the small-cluster correction, underscoring the need for replication. Mixed-effects trajectory analyses were directionally consistent with the lagged findings, but baseline IGF-1 did not improve the better-fitting random-slope model or clearly account for the substantial between-participant heterogeneity in longer-term progression rates. Exploratory analyses did not establish a nonlinear threshold, demonstrate variation across observed progression categories, or show improved classification of clinically meaningful decline. These findings position circulating IGF-1 as a biologically coherent candidate marker of systemic trophic-metabolic reserve rather than a dominant standalone prognostic biomarker or direct therapeutic target, supporting its further evaluation within larger longitudinal and multimodal biomarker frameworks.

## Ethics statement

Ethics approval was obtained from the Macquarie University Human Ethics Committee (Ref: 5201600387 and 5201700825).

## Data Availability

All data produced in the present study are available upon reasonable request to the authors

## Acknowledgements

Biospecimens and related clinical data used in this research were obtained from the Neurodegenerative Disease Biobank, Macquarie University, New South Wales, Australia.

## Authors contributions

**Conceptualization:** J.D.A., C.K.L.; **Methodology:** C.K.L.; **Investigation:** J.H; MB, J A. H, SF, **Data curation, analysis & visualization:** C.K.L.; **Writing—original draft:** J.D.A., C.K.L; **Writing—review and editing:** all authors; **Diagnosed and managed the care of all patients -** D.B.R **Supervision:** J.D.A. **Project administration:** J.D.A.; **Funding acquisition:** J.D.A., D.R., C.K.L. All authors read and approved of the final manuscript.

