## Supplemental Material for "Circulating insulin-like growth factor 1 (IGF-1) and short-term functional change in amyotrophic lateral sclerosis (ALS): exploratory evidence for systemic trophic-metabolic reserve"

**Running title:** IGF-1 and ALSFRS-R in ALS

Chai K Lim[1,3#], Mariana Brocardo[1], Julie Hunter[1,2], Judith A. Heads[1, 2], Dominic B Rowe[1, 2], Sarah Furlong[1, 2], Julie D Atkin[1, 2*]

Affiliations:

1. MND Research Centre, Faculty of Medicine, Health and Human Sciences, Macquarie University, Sydney, NSW 2109, Australia
2. Brain Institute and Clinic, Macquarie Medical School, Faculty of Medicine, Health and Human Sciences, Macquarie University, Sydney, NSW 2109, Australia
3. Department of Child and Adolescent Psychiatry, Psychosomatic Medicine and Psychotherapy, Jena University Hospital, Jena, 07743, Germany

#Current affiliation

* Correspondence to Prof. Julie D. Atkin

Macquarie University

75 Talavera Road, NSW 2109, Australia

Keywords:

Amyotrophic lateral sclerosis, biomarker, IGF-1, ALSFRS-R

### Supplementary Methods

#### Analysis sample and general approach

Exploratory analyses used the same complete visit-to-visit transition sample as the primary lagged analysis (48 intervals from 23 participants). The outcome was change in ALSFRS-R between consecutive visits, defined as the subsequent score minus the prior score; therefore, higher or less negative values indicated more preserved function. Unless otherwise specified, models adjusted for inter-visit interval in years, baseline BMI standardised per SD, age in years, and sex, with female as the reference category. Lagged IGF-1 denotes the concentration measured at the prior visit. Linear-model inference used participant-clustered HC1 covariance estimates to account for repeated intervals within participants. All analyses were exploratory and all tests were two-tailed.

#### Exploratory nonlinearity and threshold analyses

Potential nonlinearity in the association between prior-visit IGF-1 and subsequent ALSFRS-R change was examined using a cubic B-spline with three degrees of freedom. The joint contribution of the spline basis terms was evaluated using a participant-clustered Wald test. Adjusted predictions were generated across the observed IGF-1 range with inter-visit interval and age held at their sample means, baseline BMI held at its standardised mean of zero, and sex held at the reference or modal category. The 95% confidence band was obtained by participant-level bootstrap resampling, retaining all intervals from each resampled participant.

Complementary analyses assessed robustness to the assumed functional form. Lagged IGF-1 was modelled in tertiles, as a binary variable using the exploratory threshold of 145, and using a hinge model centred at 145 to test whether the slope changed above this value. These models used the same covariates and participant-clustered inference as the primary lagged model.

#### Effect modification by observed progression category

To examine whether the prospective association differed across observed slow, normal, and fast progression categories, a model including lagged IGF-1, progression category, and their interaction was fitted. The interaction terms were assessed jointly using a participant-clustered Wald test. Because progression category summarised the observed disease course, this analysis was treated as exploratory.

#### Incremental explanatory value of IGF-1

A base linear model containing observed progression category, inter-visit interval, standardised baseline BMI, age, and sex was compared with an extended model additionally containing standardised lagged IGF-1. The incremental contribution of IGF-1 was summarised using the change in ordinary $R^2$ and partial $R^2$. The coefficient and p-value for lagged IGF-1 in the extended model were calculated using participant-clustered covariance estimates.

**Classification of clinically meaningful decline**

Clinically meaningful decline was defined as a decrease of at least 5 ALSFRS-R points between consecutive visits. Ridge-penalised logistic regression was used because of the modest number of events relative to the number of predictors. The base model included observed progression category, inter-visit interval, baseline BMI, age, and sex; the extended model additionally included lagged IGF-1. Discrimination was summarised using the area under the receiver operating characteristic curve (AUC). Apparent AUC was calculated in the complete analysis sample. Internal performance was then assessed using five-fold stratified participant-grouped cross-validation, ensuring that all intervals from a participant were retained entirely within either the training or test set in each fold. Continuous predictors were standardised within the training data and the same transformations were applied to the corresponding test data. Pooled out-of-fold predictions were used to calculate the cross-validated AUC.

### Supplementary Results

#### Exploratory nonlinearity and threshold analyses

The adjusted spline suggested a possible nonlinear pattern in which lower prior-visit IGF-1 concentrations corresponded to greater subsequent functional decline, with apparent flattening at higher concentrations (**Figure S1**). However, the spline terms were not jointly significant (overall $p = 0.190$). Complementary analyses also did not establish a reproducible nonlinear relationship: the overall tertile test was not significant ($p = 0.179$), participants above the exploratory threshold of 145 had 2.30 points less decline than those below it but this contrast was not statistically significant ($p = 0.115$), and the hinge term did not support a change in slope above 145 ($p = 0.592$). These findings therefore do not establish either a specific functional form or a clinically meaningful IGF-1 threshold.

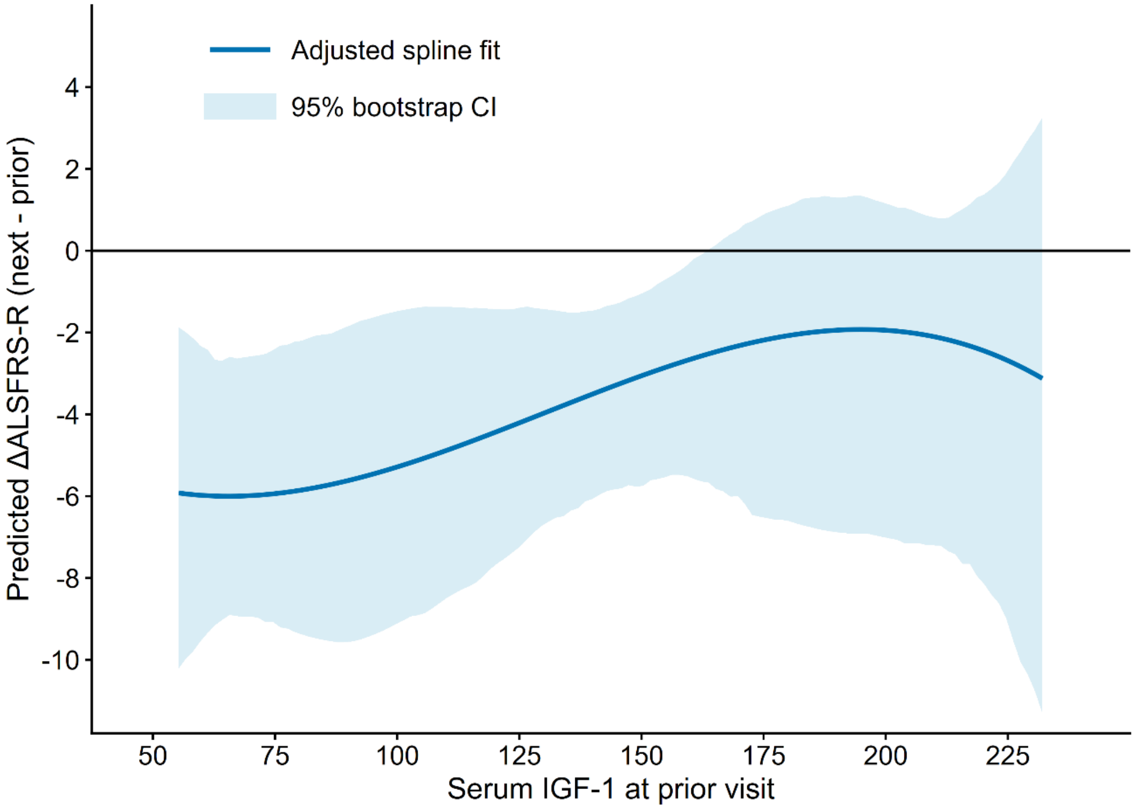


***Figure S1. Exploratory nonlinear association between lagged IGF-1 and subsequent ALSFRS-R change.*** *Predicted ΔALSFRS-R, defined as the subsequent visit minus the prior visit, is shown as a function of serum IGF-1 at the prior visit. The solid line represents the adjusted cubic B-spline fit controlling for inter-visit interval, standardised baseline BMI, age, and sex. The shaded area denotes the 95% participant-cluster bootstrap confidence interval.*

#### Effect modification by observed progression category

There was no evidence that the association between prior-visit IGF-1 and subsequent ALSFRS-R change differed across observed progression categories (joint IGF-1 × progression-category interaction, $p = 0.995$). The estimated short-term association was therefore broadly similar across the slow, normal, and fast progression categories, although the modest subgroup sizes limited precision.

#### Incremental explanatory value of IGF-1

Adding lagged IGF-1 to the model containing observed progression category, inter-visit interval, baseline BMI, age, and sex increased $R^2$ from 0.369 to 0.383 ($\Delta R^2 = 0.014$; partial $R^2 = 0.022$). However, the incremental IGF-1 coefficient was not statistically significant using participant-clustered inference ($p = 0.183$), indicating that the additional explanatory contribution was modest and estimated with considerable uncertainty.

#### Classification of clinically meaningful decline

At the prespecified threshold of at least 5 ALSFRS-R points of decline, 17 of 48 intervals were classified as events. The apparent AUC was 0.834 for the clinical model and 0.824 after adding lagged IGF-1 ($\Delta$AUC = − 0.010). Participant-grouped cross-validation yielded AUCs of 0.6973 and 0.6755, respectively ($\Delta$AUC = − 0.0218). Thus, adding IGF-1 did not improve either apparent or internally cross-validated discrimination of clinically meaningful decline.

#### Sensitivity across alternative decline thresholds

Classification analyses repeated across ALSFRS-R decline thresholds of 3 – 9 points showed no consistent improvement after adding IGF-1. Small positive changes in AUC at some of the more severe thresholds were unstable, with confidence intervals that included no improvement, and were based on small event counts. These threshold-sensitivity analyses were therefore considered hypothesis-generating.

***Table S1. Summary of exploratory analyses***

| Analysis | Comparison / estimate | Statistical result | Interpretation |
|---|---|---|---|
| Cubic B-spline | Joint spline terms | $p = 0.190$ | Allowing a flexible nonlinear IGF-1 association did not clearly improve the model |
| IGF-1 tertiles | Overall category test | $p = 0.179$ | Subsequent ALSFRS-R change did not clearly differ across low, middle, and high IGF-1 groups |
| Threshold at 145 | Above vs below 2.30 ALSFRS-R points | $p = 0.115$ | Participants above 145 showed numerically less decline, but the difference was uncertain |
| Hinge model at 145 | Change in slope above 145 | $p = 0.592$ | No evidence that the IGF-1 association changed after 145 |

| Analysis | Comparison / estimate | Statistical result | Interpretation |
|---|---|---|---|
| Progression interaction | IGF-1 × progression category | $p = 0.995$ | The IGF-1 association was similar across slow, normal, and fast progressors |
| Incremental explanatory value | $R^2$ 0.369 – 0.383; $\Delta R^2 = 0.014$; partial $R^2 = 0.022$ | $p = 0.183$ | IGF-1 explained only a small amount of additional variance, with uncertain improvement |
| Apparent classification | AUC 0.834 – 0.824 | ΔAUC = −0.010 | Adding IGF-1 slightly reduced in-sample discrimination |
| Participant-grouped cross-validation | AUC 0.6973 – 0.6755 | ΔAUC = −0.0218 | Adding IGF-1 reduced discrimination when tested in held-out participants |

***Note.*** *All linear-model p-values use participant-clustered HC1 covariance estimates. AUC, area under the receiver operating characteristic curve; BMI, body mass index; IGF-1, insulin-like growth factor 1.*
